# Yield of genetic association signals from genomes, exomes, and imputation in the UK biobank

**DOI:** 10.1101/2023.09.13.23295479

**Authors:** Sheila M. Gaynor, Tyler Joseph, Xiaodong Bai, Olga Krasheninina, Boris Boutkov, Evan Maxwell, Suganthi Balasubramanian, Anthony Marcketta, Joshua Backman, Regeneron Genetics Center, Jeffrey G. Reid, John D. Overton, Luca A. Lotta, Jonathan Marchini, William J. Salerno, Aris Baras, Goncalo R. Abecasis, Timothy A. Thornton

## Abstract

Whole genome sequencing (WGS), whole exome sequencing (WES), and array genotyping with imputation (IMP) are common strategies for assessing genetic variation and its association with medically relevant phenotypes. To date there has been no systematic empirical assessment of the yield of these approaches when applied to 100,000s of samples to enable discovery of complex trait genetic signals. Using data for 100 complex traits in 149,195 individuals in the UK Biobank, we systematically compare the relative yield of these strategies in genetic association studies. We find that WGS and WES combined with arrays and imputation (WES+IMP) have the largest association yield. While WGS results in a ∼5-fold increase in the total number of assayed variants over WES+IMP, the number of detected signals differed by only 1% for both single-variant and gene-based association analyses. Since WES+IMP typically results in savings of lab and computational time and resources expended per sample, we evaluate the potential benefits of applying WES+IMP to larger samples. When we extend our WES+IMP analyses to 468,169 UK Biobank individuals, we observe a ∼4-fold increase in association signals with the ∼3-fold increase in sample size. We conclude that prioritizing WES+IMP and large sample sizes, rather than current short-read WGS alternatives, will maximize the number of discoveries in genetic association studies.

## Introduction

Large-scale genetic studies provide insight into the biological underpinnings of a wide range of human traits, guiding the development of new therapeutic interventions and disease prevention strategies through improved understanding of human health and disease. Examples of new therapies enabled by genetic studies include blocking *PCSK9* for the prevention of recurrent heart disease [1-3], blocking *ANGPTL3* for the treatment of familial hypercholesterolemia [4], and CRISPR editing of *BCL11A* for the treatment of sickle cell disease [5, 6]. Genetic association studies can now include extensive health data across millions of participants and have a choice of diverse approaches for capturing genetic variation, ranging from array genotyping followed by imputation (IMP) to whole exome sequencing (WES) to whole genome sequencing (WGS). Each of these approaches differ in the number and type of variants captured, the fidelity of information provided, and cost and complexity of execution. However, the impact of choosing each approach on detection of actionable genetic association signals remains unclear.

For the past 15 years, genotyping and imputation has been the workhorse of genetic association studies [7, 8]. Studies based upon array genotyping followed by continually improved imputation are routinely applied to complex traits ranging from macular degeneration [8], to inflammatory bowel disease [9, 10], to schizophrenia [11] and have produced 10,000s of genetic association signals [12]. Array genotyping and imputation enables the study of relatively common variants (typically, those with frequencies >0.1 – 1% and which have been characterized in a reference set of individuals) [13]. Recent examples include studies of COVID susceptibility in hundreds of thousands of individuals, identifying genetic variants that lower *ACE2* expression as protective factors, and identifying variants in *IFNRA2* and other immune related genes as major determinants of susceptibility [14, 15]. Nonetheless, translating GWAS findings into actionable insights for biological understanding and therapeutic intervention remains a laborious and ongoing process [16]. These challenges in translation arise because most GWAS findings point to non-coding variants, variants whose function is uncertain, and/or variants with small effect sizes.

In the past 10 years, exome sequencing has emerged as a practical and successful strategy for uncovering the genetic basis of human disease. WES captures protein-coding variants as rare as singletons that are beyond the reach of arrays and imputation. Exome sequencing was originally used to identify the causes of hundreds of rare Mendelian disorders, by studying collections of individual cases and their families [17]. Exome sequencing studies of complex traits in large population samples are increasingly common. They have yielded rare coding variant association signals that are easier to interpret and experimentally follow-up than those found in GWAS. Translation for these signals is easier when variants have large effect sizes and connect impaired function of a specific gene to a therapeutic outcome. Recent examples include studies of rare genetic variants that protect against obesity and liver disease [18, 19], pointing to *GPR75* and *CIDEB* as potential therapeutic targets. WES and IMP are often used together as an effective approach for capturing both common variants genome-wide and rare protein-altering variants in coding regions.

Most recently, short-read whole genome sequencing has been applied at scale to dissect Mendelian diseases [20] and common diseases [21-23]. WGS studies aim to capture coding and non-coding variation across the allele frequency spectrum and much larger numbers of genetic variants than either IMP or WES based approaches. There is widespread excitement about its potential for enabling genetic discovery [24, 25]. WGS has the most substantial costs, including data generation, processing, storage, and analysis, but these are decreasing [26]. At the present time it remains unknown whether deployment of WGS at scale will enable a wave of genetic discoveries comparable to those that resulted from the deployment of arrays, imputation, and whole exome sequencing.

Understanding the relative performance of these approaches is essential for the design of current and future genetic studies. Here, we systematically assess variants captured and the resulting discovery yield for genetic association studies in large biobank samples using IMP, WES, and WGS. For these comparisons, we rely on data from 149,195 UKB participants that have been characterized with each of these assays and for whom extensive health information is available. We first perform a survey of the genetic variants that can be assessed with each approach and then compare the yield of single variant and gene-based association signals across a set of 100 traits in this large UKB sample. Finally, recognizing that choice of analysis approach could entail changes in sample size, we evaluate association yield when varying the sample size using the same set of 100 traits. Our study provides an empirical assessment of these approaches and enables investigators to make informed decisions about large-scale genetic studies.

### UK Biobank Dataset: Genomes, Exomes and Imputation

Our primary analyses included 149,195 UK Biobank participants with data from IMP [27], WES [28, 29], and WGS [21] as described in Supplementary Figures 1-2 and Supplementary Table 1. The UKB used a custom designed SNP array with 805,426 variants designed to capture common variation and selected high-value protein altering variants. We extended this data to an additional 111,333,957 variants through imputation using TOPMed Freeze 8 genomes as a reference panel [22, 30, 31]. WES targeted the coding region of 18,893 genes (∼1% of the genome) and sequenced 95% of these bases to a depth of >25X in each individual [28, 29]. WES resulted in 17,131,8674 variants, of which 10,463,945 were in coding regions. Finally, WGS was carried out to a depth of >20X using short-read sequencing, ensuring capture of > 95% of the genome with >15X depth. Overall, WGS resulted in 599,385,545 variants [21]. We applied uniform processing and filtering to the variant sets from each approach (see Methods for details; key filtering steps included removing variants known to have failed in prior large-scale WGS efforts). We evaluated 4 different strategies: IMP, WES, WGS and WES+IMP; full results for each strategy are included in tables, figures and supplementary information but for ease of presentation we focus on the comparison of WES+IMP and WGS, which were the two most powerful approaches.

Overall, the WGS strategy identified 599,385,545 SNVs and indels whereas the WES+IMP strategy identified 125,694,205 SNVs and indels (Fig. 1, Panels A-B). Variants jointly captured by each of the platforms were 99.9% concordant (Supplementary Table 2). In contrast to the total number of variants, the count of variants observed in each individual was similar between the two approaches: mean: 3,595,704 variants per individual in WGS vs 3,585,289 in WES+IMP (Fig. 1, Panel C). The bulk of variants unique to WGS are very rare and present in only a few individuals each, explaining how a ∼5-fold increase in overall variant count translates into ∼0.3% more variants per individual (Fig. 1, Panel D). For example, 47% of WGS variants are singleton variants present in one individual, whereas only 7% of WES+IMP variants are singletons.

**Figure 1.**
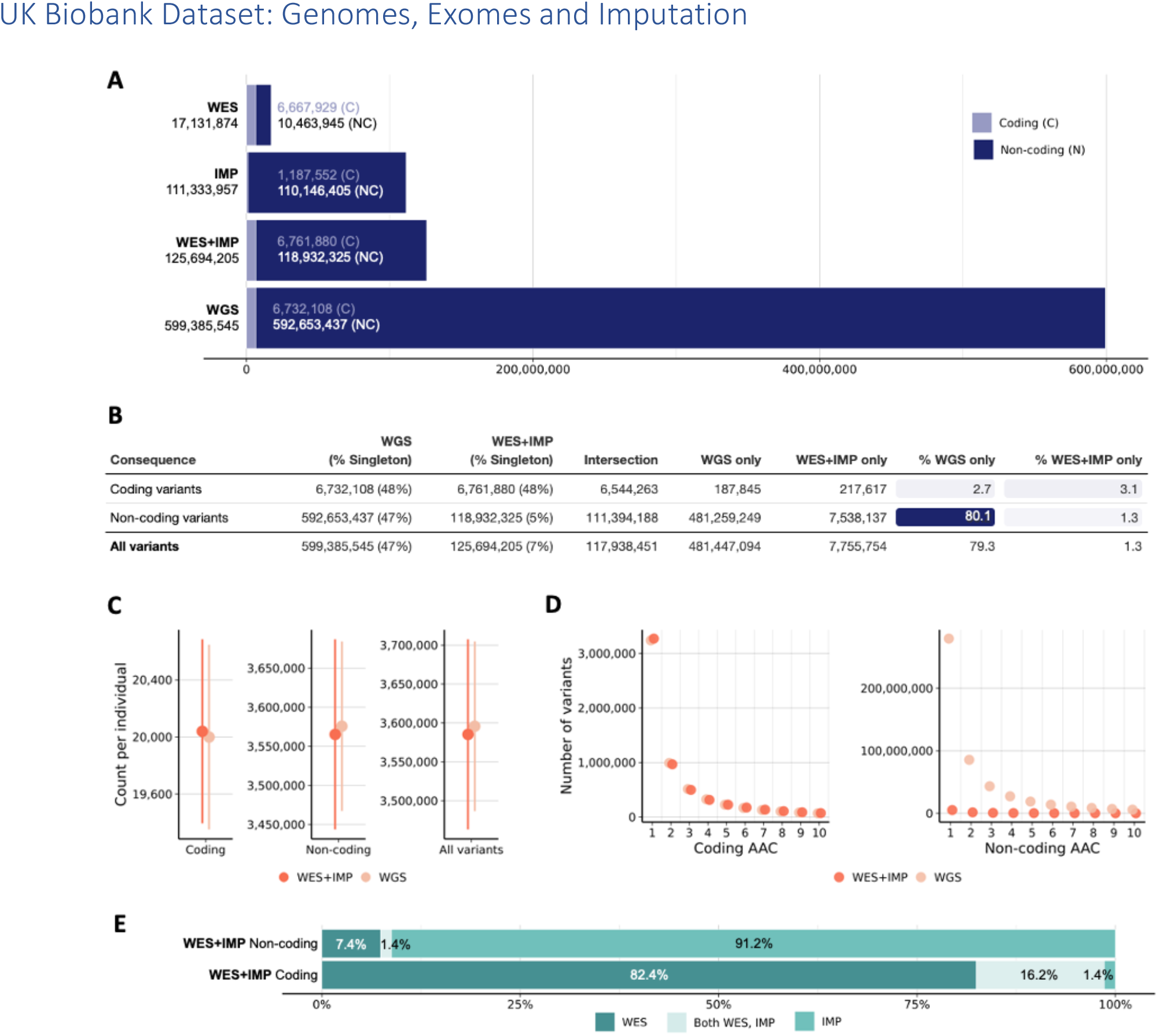
a. The number of coding and non-coding variants for all variants in the WES, IMP, combined WES+IMP, and WGS datasets. b. A comparison of variants observed by the WES+IMP and WGS datasets by variant consequence, with the relative gains in approach-exclusive variants. c. The number of variants observed per individual; the point provides the average and the error bars representing the corresponding standard errors. d. The number of non-coding and coding variants observed at the lowest alternative allele counts for the WES+IMP and WGS data. e. The percentages of non-coding and coding variants observed only in WES, in both WES and IMP, and only in IMP in the combined WES+IMP dataset.

Variant identification in coding regions was very similar: the total number of coding variants was 6,732,108 variants for WGS versus 6,761,880 variants for WES+IMP; and the mean count of coding variants per individual was 20,000 for WGS versus 20,039 for WES+IMP (Fig. 1, Panel C). Within coding regions, 48% of variants were singletons for both WGS and WES+IMP. The differential contributions of WES and IMP to the coding and non-coding variant sets in WES+IMP are described in Fig 1, Panel E. Coding variation is further described in the Supplementary Information, Supplementary Figure 3, and Supplementary Tables 3-4.

### Single Variant Tests

We assessed differences in genetic association yield in two stages. First, we evaluated differences in yield for single variant tests (which are responsible for the bulk of known genetic association signals). Second, we evaluated differences in yield in gene-based tests (discussed in the next section; Supplementary Figure 4), which can provide more specific insight about the underlying biology. We performed association tests across 80 quantitative and 20 binary traits and first tested each variant present in at least 5 individuals for association using REGENIE [32] (see Methods for further details). All tests are summarized in Fig. 2.

**Figure 2.**
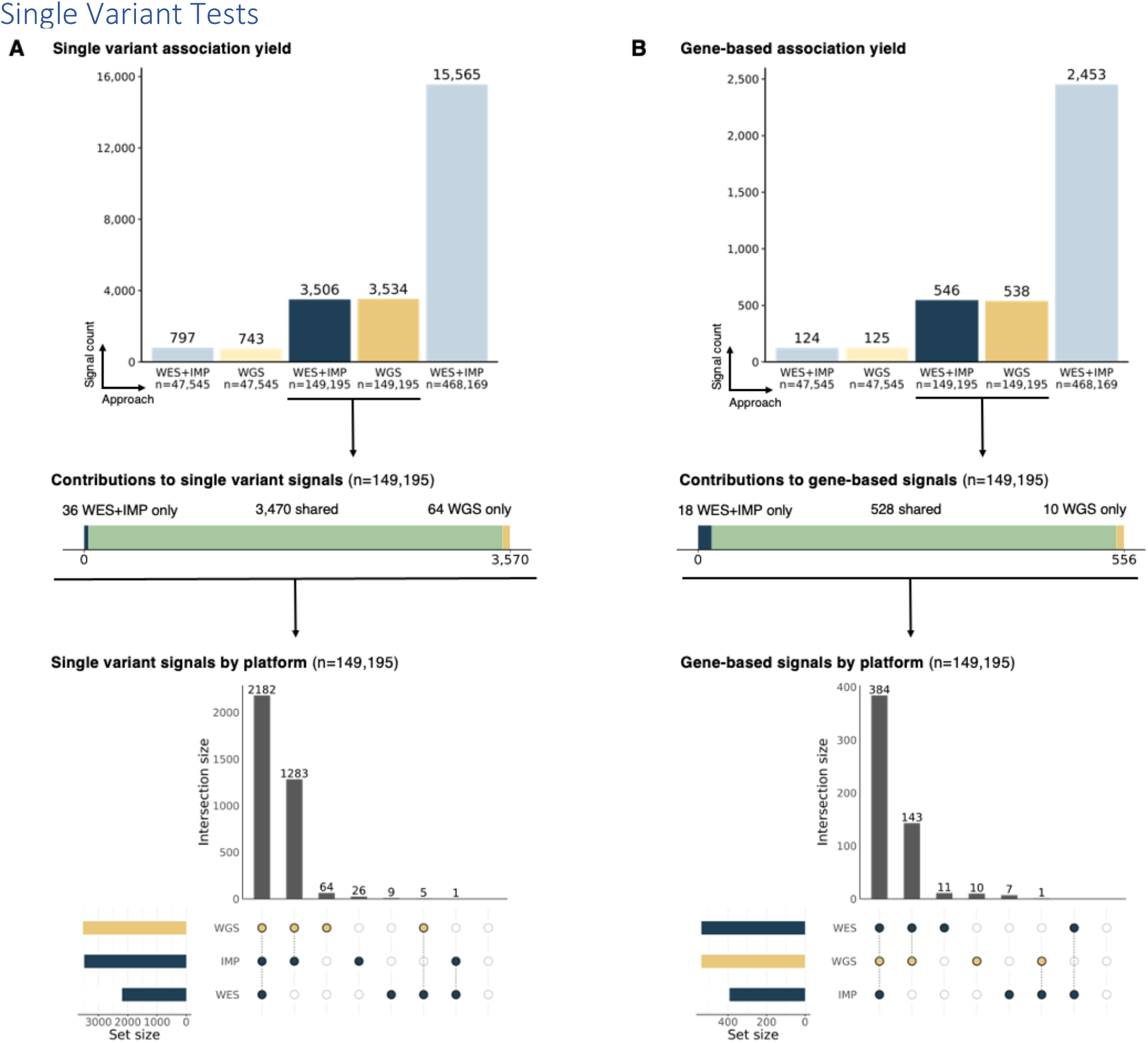
a. Single variant association signals identified across each analysis sample, where platform-specific results are specified for the primary analytical sample n=149,195. b. Gene-based association signals identified across each analysis sample, where platform-specific results are specified for the primary analytical sample n=149,195.

We identified 3,570 genome-wide significant signals across all trait-locus pair combinations (Supplementary Table 7). We found a similar number of signals from WES+IMP (3,506 or ∼35.1 per trait) and from WGS (3,534 or ∼35.3 per trait, a 1% increase). Nearly all signals were found using both approaches (3,470 of 3,570, or 97.2%) and most of these pointed to the exact same variant (96%). Supplementary Figure 5 compares frequencies, effect sizes, and p-values across signals; Supplementary Table 5 summarizes annotated consequence of the peak variant for each signal. Across all platforms, the same highly significant associations with the largest -log_10_ p-value typically corresponded to high allele frequency variants and small effect sizes. Shared signals were very comparable beyond the peak variant (Supplementary Figure 6 summarizes a typical shared signal across platforms). Such shared associations included signals in genes used to inform therapeutics; for instance, the peak signal in *PCSK9* with LDL cholesterol of missense variant 1:55039974:G:T (AAF=0.018) was identified across WGS (P=4.18 × 10^−140^), WES (P=4.18 × 10^−140^), and IMP (P=6.23 × 10^−140^).

In contrast to the shared signals, nearly all signals found with WGS only (64 signals) or WES+IMP only (36 signals, were relatively marginal in p-value (60% within two orders of magnitude of the significance threshold), pointed to rare variants with frequency less than 0.1% (67%) and typically had limited support from nearby variants. Supplementary Figures 7-9 show examples of signals specific to each platform. We sought to replicate our findings using WES+IMP data from the remaining UKB individuals (n=318,974). We replicated 17 of 36 signals only in WES+IMP, 13 of 64 signals only in WGS, and 3,386 of the 3,570 shared signals. A large-scale WGS dataset for replication of WGS only signals is unavailable at present. We interpret the observation that platform specific signals are relatively weak, led by rare variants, lack support from nearby variants, and failed to replicate as evidence that many of these are artifacts.

### Gene-Based Tests

In contemporary genetic association studies, gene-based tests – which aggregate evidence across rare protein altering variants in a gene – provide a powerful complement to single variant tests and are especially important for identifying biological insights. These tests directly point to biological effector mechanisms and can often identify sets of variants with very large effects [28, 29]. We thus followed up our analysis of single variants with a series of gene-based tests. We examined coding variants in each gene grouped according to frequency (AAF < 1%, <0.1%, <0.01%, <0.001% and singletons) and functional consequence (missense, deleterious missense, and pLOFs) and considered association tests where all variants in a gene act in the same or in different directions (see Methods, Supplementary Table 6). We summarized all association tests for gene-trait pairs using GENE_P [33], a single, unified p-value (Methods), and found similar results when examining individual tests or frequency thresholds.

We observed associations between 556 gene-trait pairs across the WES+IMP and WGS analysis (Supplementary Table 8). We again found a similar number of signals from WES+IMP (546) and from WGS (538, a 1% decrease). Consistent with the single-variant analysis, 95% of gene-based association signals (528 of 556 unique signals) were identified by both WES+IMP and WGS. Again, associations with the largest -log_10_ p-value were consistently observed in WGS and WES+IMP (Supplementary Figures 10-11). The shared signals included several known true positives, including again therapeutically relevant genes like the *PCSK9* association with LDL cholesterol (WGS P=1.27 × 10^−80^, WES P=3.91 × 10^−82^, and IMP P=1.42 × 10^−36^; Supplementary Table 8).

When we examined the 10 signals exclusive to WGS and the 18 signals exclusive to WES+IMP, we found that half were signals where one approach exhibited subthreshold association (2.6 × 10^−7^< P <1 × 10^−5^) and the addition of a small number of approach-specific variants increased signal to significance for the alternate approach. The remaining cases were driven by a single variant that was platform-exclusive (1 variant in CALR for WGS, 1 variant in OMA1 in WES, 6 different variants in IMP). This contrasts with the single variant results, which appeared more enriched for platform-specific artifacts. We again sought to replicate these signals in the remaining UKB samples, and replicated 14 of 18 signals exclusive to WES+IMP, 5 of 10 signals exclusive to WGS, and 514 of the 556 shared signals.

### The Importance of Sample Size

Advancing our understanding of human health and disease requires balancing the returns from each technology when applied to the same sample but also the consequences of examining different numbers of samples. To assess the impact of sample size on association study yield, we repeated all analyses on a subset of n=47,545 individuals in UKB with WGS and WES+IMP and on the entire n=468,169 UKB sample with WES+IMP data (Supplementary Figures 1-2, Supplementary Table 1), which correspond to ∼3-fold changes in sample size relative to our primary analysis with n=149,195 individuals.

When we focused our analyses on the set of 47,545 individuals (a three-fold decrease in sample size), the number of single variant signals decreased more than four-fold from 3,534 to 743 for WGS and from 3,506 to 797 for WES+IMP and the number of gene-based signals decreased more than four-fold from 538 to only 125 for WGS and from 546 to 124 for WES+IMP. When we extended the WES+IMP analysis from 149,195 individuals 468,169 individuals (a three-fold increase in sample size), the number of single variant signals increased more than four-fold from 3,506 to 15,565. Similarly, the number of gene-based association signals also increased four-fold from 546 to 2,453. Larger sample sizes permitted the identification of additional rare variant signals, often for variants present in the smaller dataset but with insufficient power. Comparing the analyses of 47,545 individuals to the analyses of 468,169 individuals, we find the ∼10-fold increase in sample size resulted in a ∼20-fold increase in association yield.

Thus, whereas choosing between WGS and WES+IMP changed the number of signals by about 1%, changes in experimental design that enable larger samples through lab efficiencies, cost, and analytical simplicity can have a much greater impact on discovery yield. For example, whereas WES+IMP for all UKB samples has been publicly available since 2021, the complete WGS data is still not publicly available as of this writing.

### Signals from each approach alone

Our analyses thus far have shown that the combination of WES+IMP is equivalent to WGS for practical purposes when focused on association yield. It can also be important to consider the relative yield from WES alone and IMP alone.

For single variant association signals, we found IMP and WGS to be roughly equivalent. Nearly all the variants that could drive a significant association signal by themselves were common enough to be imputed. Even though 90% of peak variants were non-coding (Supplementary Table 5), WES alone was able to find 62% of single variant association signals. WES was able to find these signals because common variant association signals are enriched near genes and because, through linkage disequilibrium, most single variant association signals are supported by nearby variants. However, using WES to detect non-coding single variant signals resulted in substantial loss of fidelity, often identifying different peak variants than WGS and IMP (which agreed 97% of the time).

Broadly speaking, WES and WGS found the same sets of gene-based association results, whereas IMP alone missed 29% of the gene-based signals. Among all significant gene-based signals, the contributions of very rare variants cannot be identified by IMP alone due to limited capture at the lowest frequencies. For example, among all significant signals there were 81 gene-trait pairs where WGS and/or WES found association between a burden of singleton coding variants (whether pLOF or missense) and a trait at p < 2.6 × 10^−8^. None of those singleton signals could be found through imputation alone but 38% could be identified across all burden tests in IMP. In terms of mechanistic insight, these singleton signals are among the most compelling – they refer to loci where a group of individuals, each with a unique defect in the same gene, shows a significant change in phenotype. Signals driven by rare coding variation also corresponded to some the largest effect association signals. Thus, while WGS and WES found quantitative trait signals associated with a change in trait values of >1 standard deviation for 71 and 68 gene-trait pairs respectively, IMP found only 15 such signals. For binary traits, there were 8 signals each in WGS and WES significantly associated with an increase or decrease in disease risk of >2x, but IMP found only two such signals. For example in the *GCK* gene, rare pLOF and deleterious missense variants with frequency of < 0.0001 are strongly associated with type 2 diabetes in WES (OR = 10.0, P<10^−20^) and WGS (OR = 10.1, P<10^−22^), but not in IMP (all p>0.01).

## Discussion

Genetic association studies can elucidate human biology as well as support and guide the development of new, life-changing therapies that improve population health [37]. Understanding the different strategies for maximizing the yield of these studies, particularly when measured as disease-associated signals with clear mechanistic implications, is critical to enable the best use of limited resources. Our analyses of WGS versus WES+IMP in the same UKB individuals show that WGS captures many more genetic variants but changes the numbers of association signals only modestly. WGS increased the total number of variants by >350% (from 125,694,205 to 599,385,545) and changed the number of association signals by 1% (from 3,506 to 3,534 for single variants and 546 to 538 for gene-based tests).

The cost and complexity of biobank-scale WGS is substantial compared to WES+IMP and in our analysis results in 1% higher yield for single variants and 1% lower yield for gene-based tests. Optimal use of resources must consider the value of the genetic data on additional samples or deeper phenotypic characterization (perhaps proteomics and RNA-sequencing) of the same samples as alternatives to broad WGS [38-41]. Our results show that increasing sample size is an extremely effective strategy for improving discovery yield. For example, a ∼3-fold increase in sample size resulted in a ∼4-fold increase in signals, and a ∼10-fold increase in sample size resulted in a ∼20-fold increase in signals.

The limited additional association yield from WGS is likely because most WGS-only variants are too rare to detect unless they are grouped with variants of similar function, which can presently only be done in coding regions. To date, grouping rare non-coding variants for analysis remains a major challenge, which may be informed by ongoing improvements in employing omics assays at scale. When we attempted sliding window or enhancer-based grouping (data not shown), variant aggregation outside the coding regions did not produce additional signals. Thus, while coding variants have been effectively grouped into aggregated sets to analyze the lowest frequency variants, our ability to classify, group, and interpret non-coding variants is not yet advanced enough to similarly analyze the rarest non-coding variants and increase discovery yield.

Each of the approaches we used can be used to capture variants beyond SNPs and indels. For instance, when we analyzed 637,039 structural variants (median: 7,952 structural variants per individual) detected in the WGS data, it resulted in only 16 additional association signals (3,586 with SVs and 3,570 without SVs) beyond those captured from SNVs and indels across the 100 traits examined. As sequencing approaches evolve beyond the current short-read standards, it may be possible for variants beyond SNPs and indels to make bigger contributions to discovery.

Based on our large-scale analyses of IMP, WES, and WGS, it is our view that optimizing genetic discovery currently benefits from allocating resources towards characterizing the largest possible sample sizes with WES+IMP. Resources saved from not deploying contemporary short-read WGS in the largest sample sets can be used for population-scale functional assays that will help understand how to annotate non-coding variation. At the same time, we expect that as more targeted samples are sequenced with WGS, the performance of imputation-based approaches will continue to increase and thereby further decrease any potential relative advantage of biobank-scale WGS. Thus, WGS in studies such as the Trans-Omics for Precision Medicine Program, the Mexico City Prospective Study, and in the UK Biobank Program [13, 22, 42] may provide limited benefits through novel discoveries but will improve the quality of other ongoing genetic studies through improved imputation. In our view, until WGS costs decrease substantially, the greatest benefit of further WGS for genetic association discovery will be to further increase the diversity and availability of imputation resources. Our analyses demonstrate the extent of associations identified by increasingly large-scale analyses, where cost savings from limiting WGS can be used to enable the biological follow-up that is essential to translate genetic discoveries into benefits for human health.

## Supporting information

Supplementary Materials for Yield of genetic association signals from genomes, exomes, and imputation in the UK biobank

## Data Availability

All data produced in the present work are contained in the manuscript

## Methods

### Ethical approval and informed consent

The present analysis was conducted under the approved UK Biobank application 26041. The UK Biobank project has ethical approval reviewed and provided by the North West Research Ethics Committee. Informed consent was provided by all study participants.

### UKB data preparation

The sample collection and preparation for the UK Biobank was previously described for each platform: arrays [27, 43], WES [28, 29], and WGS [21]. These approaches are summarized below.

#### Array genotyping

For array genotyping, DNA was extracted and provided in aliquots to Affymetric Research Services Laboratory for genotyping. Blood samples were genotyped using the UK Biobank Axiom array with 805,426 variants. The samples were processed in 106 batches using a custom multi-batch genotype calling pipeline. Multiple quality checks including marker-based (evaluating new markers and effects based on factors such as batch, plate, sex, and array) and sample-based (missing rates and heterozygosity) were performed.

#### Exome sequencing

For exome sequencing, DNA was prepared in fragments of 200 base pairs on average with 10-base-pair unique bar codes. Samples were processed with IDT’s xGen probe library, PCR-amplified, and quantified by quantitative PCR. Multiplexed samples were sequenced with 75-base-pair paired-end reads on the Illumina NovaSeq 6000 platform with S2 or S4 flow cells. The OQFE protocol was applied for reference alignment to GRch38, variants were called using DeepVariant, aggregation was performed using GLnexus to generate a project-level pVCF, and quality checks were performed. Using this approach, 96% of targeted bases were covered at a depth of >20X.

#### Genome sequencing

A pseudo-random subset of samples was selected for genome sequencing. DNA was prepared in fragments of 450-500 base pairs on average with barcode tracking. Samples were processed with IDT for Illumina, purified, and pooled. Libraries were prepared from paired-end reads on the Illumina NovaSeq 6000 platform with S4 flow cells. The deCODe pipeline was applied for reference alignment to GRch38, processing and merging, variant calling using GATK HaplotypeCaller and Graphtyper, and quality checks were performed. In this approach, samples had an average depth of >23.5X.

### Imputation of array datasets using the TOPMed Freeze 8 reference panel

Arrays were imputed using the TOPMed Freeze 8 reference panel on the Michigan Imputation Server [30]. Array variants were selected previously [29], based on UKB imputation use with HRC and ability to liftover to GRCh38, and uploaded in randomized batches for imputation on the imputation server. The resulting VCF files were merged and concatenated, then subset to the individuals included in the present analysis. To retain high quality imputed genotypes, variants with MAC^3^5 and MACH r^23^0.3 were retained.

### Ancestry assignment

Individuals’ continental ancestry was assigned as previously described [29]. Briefly, PCs were computed using the HapMap3 samples as reference, including all SNPs shared with the UKB array data, and then each of the UKB samples were projected onto the PCs. A kernel density estimator (KDE) was trained and used to calculate the likelihood of a sample belonging to a continental ancestry group: African (AFR), Hispanic or Latin American (HLA), East Asian (EAS), European (EUR), and South Asian (SAS). Samples with a likelihood of a single ancestry greater than 0.3 were assigned that ancestry; samples with a likelihood of two ancestries greater than 0.3 were assigned AFR over EUR, HLA over EUR, HLA over EAS, SAS over EUR, and HLA over AFR. If ancestry likelihoods were all less than 0.3 or three ancestries were greater than 0.3, the sample was excluded from analysis.

### Analysis-ready dataset preparation

#### Sample selection

To permit comparison across platforms, we identified all individuals with array genotyping, exome sequencing, and genome sequencing available. This yielded a sample of n=149,195 and was our primary analytical set. To evaluate the influence of sample size, we also constructed a sample from all individuals with both array genotyping and exome sequencing. This yielded a sample of n=468,169 individuals. We also generated a sample of individuals with genome sequencing, by down sampling to retain the same multiplier (3.138) between sample sets. We randomly selected n=47,545 individuals with genome sequencing data for the final sample. Lastly, we generated a replication sample by selecting all n=318,974 individuals with WES+IMP that were not included in the primary analytical set of n=149,195 individuals.

#### Genetic data

Data was prepared for analysis within each platform using consistent filtering. For the sequencing datasets, all variants with MAC^3^1 were considered. Across all genetic datasets, we excluded variants with Hardy-Weinberg equilibrium (HWE) test P > 1 × 10^−15^ and >10% missingness. We further excluded all variants that failed QC in TOPMed Freeze 8 [22] or gnomAD [44] and failed QC as given by the GraphTyper HQ definition in Halldorsson et al [21]. Analysis-ready datasets were generated in Plink2 file sets (PGEN format).

#### Phenotype data

For the phenotype data, we used trait data from the UK Biobank Data Showcase. A set of 100 traits were selected to permit generalizable conclusions and maintain feasibility of analysis. Traits were selected from 492 traits that were previously described and identified to have rare variant associations [29]. The trait set was reduced to retain 80 quantitative and 20 binary traits by pruning for redundancy, sufficient case counts, and prioritizing trait heritability. Each of the quantitative traits were rank-based inverse-normal transformed prior to analysis.

### Gene and variant annotation

Variants were annotated using VEP [45] based upon the canonical transcript of protein-coding transcripts. Gene regions were defined using Ensembl release 100, and canonical transcripts were defined using MANE tags where available, followed by APPRIS or Ensembl tags as necessitated. Variants were defined as predicted loss of function (pLoF) when annotated as frameshift, stop gained, splice donor or acceptor, start lost, or stop lost. Missense variants were further assigned a deleteriousness score ranging from 0-5 based on five algorithms in dbNSPF [46]: SIFT [47], PolyPhen2 HDIV and HVAR [48], LRT [49], and MutationTaster [50]. Deleterious scores were grouped as “likely deleterious” when predicted deleterious by 5 of 5 algorithms, “possibly deleterious” when predicted deleterious by at least 1 of 5 algorithms, and “likely benign” when predicted deleterious by 0 of 5 algorithms.

### Association analyses

All association analyses were performed using REGENIE [32]. For each of the 100 traits, we ran Step 1 of REGENIE using the observed genotyping array data. Array variants were included with <10% missingness and HWE test *P* > 10^−15^. The resulting predictors were included as covariates in Step 2 of REGENIE, in addition to age, age squared, sex, age-by-sex, and 10 ancestry-informative PCs derived from the exome sequencing data. Our association analyses considered autosomes and chromosome X.

#### Single variant associations

For single variant analysis, each variant with MAC≥5 was tested using Step 2 of REGENIE separately for each platform. We considered a significance threshold of P=5 × 10^−12^ based on Bonferroni correction for testing 100 phenotypes genome-wide. To identify independent signals, we performed peak-finding jointly across each dataset. This approach merged all sets of results (WGS, WES, imputed array) and identified independent peaks by scanning across the genome and identifying significant associations, then selecting the most significant signal and pruning signals to only permit one significant signal per 1000 Kb. To account for shared signal obscured by the significance threshold, we considered signals to be shared when a variant has p-value within one order of magnitude of the significance threshold (P=5 × 10^−11^) in the comparative platform; platform-specific unique signals did not have any signals within one order of magnitude of significance within 1000 Kb in the comparative platform. We excluded outlier variants with high overall genotype mismatch with respect to a given platform where comparison was available across the three platforms. Replication was performed in the exact same approach for the replication sample (in WES, imputed array data), maintaining the significance threshold of P=5 × 10^−12^.

#### Gene-based associations

We performed aggregation tests on rare variants using Step 2 of REGENIE separately for each platform to identify gene-based associations in coding regions. Annotations were used to generate gene-sets by collapsing variants within gene regions based on allele frequency and variant consequence. We considered 7 consequence-based masks (Supplementary Table 7): putative loss of function (pLoF), pLoF + likely deleterious missense, pLoF + likely/possibly deleterious missense, pLoF + all missense, likely deleterious missense, likely/possibly deleterious missense, and all missense. We considered five allele frequency bins for each of the consequence-based masks, based on the alternative allele frequency thresholds: MAF≤ 1%, MAF≤ 0.5%, MAF≤ 0.1%, MAF≤ 0.01%, and singletons only. Collectively, up to 35 mask combinations were tested for each gene where sufficient data was available for testing. A unified test was performed for each gene to yield a gene-level p-value that aggregated each mask combination across all testing frameworks (burden, SKAT, and ACAT tests). We considered a significance threshold of P=2.6 × 10^−8^ based on a Bonferroni correction for all gene-based tests within each platform; signals were defined as shared if the comparative platform had p-value within one order of magnitude of significance (2.6 × 10^−7^). Platform-specific unique gene-based signals did not have P<2.6 × 10^−7^ in the comparative platform. We excluded sets with outlier variants with high heterozygous genotype mismatch, given variant rarity, with respect to a given platform where comparison was available across the three platforms. Replication was performed in the exact same approach for the replication sample (in WES, imputed array data), maintaining the significance threshold of P=2.6 × 10^−8^.

