## Supplementary Materials for Yield of genetic association signals from genomes, exomes, and imputation in the UK biobank for "Yield of genetic association signals from genomes, exomes, and imputation in the UK biobank"

### Supplementary Information

#### Survey of coding variation

Before proceeding to genetic association analyses, we compared the number of coding variants detected by the WGS and WES+IMP approaches. We annotated variants by functional consequence with the ENSEMBL Variant Effect Predictor (McLaren et al., 2016) using the ENSEMBL 100 canonical transcript definitions (Cunningham et al., 2022). As expected, both approaches resulted in very similar numbers of coding variants per individual (WGS median: 19,905, IQR: 239; WES+IMP median: 19,948, IQR: 245). For both datasets, 48% of observed variants were singletons. For WGS, 75.3% variants are present in less than 5 individuals and similarly 74.7% of WES+IMP variants are present in less than 5 individuals. Overall, coding variants were distributed across 19,377 genes in the WGS data, and across 18,446 genes in the WES+IMP data set (among the genes in WES+IMP dataset, variants in 347 genes were outside the exome target regions and detected only through arrays and imputation).

The total number of coding variants captured by each approach was very similar (WGS 6,732,108 variants; WES+IMP 6,761,880 variants) with 6,544,263 observed in both WGS and WES+IMP. Among variants that were present in only the WES+IMP dataset, there were 126,319 missense variants – compared to 88,448 missense variants specific to the WGS data – the largest increase for a coding variant consequence. In contrast, the largest proportional gain was for variants that were present only in the WGS data for in-frame indels or putative-loss-of-function (pLOF) variants – there were 9.3% more pLOFs and 23.5% more in-frame indels specific to the WGS data, but only 7.2% more pLOFs and 5.6% more in-frame indels specific to the WES+IMP data. Overall, 2.7% of coding variants were observed only in WGS and 3.1% of variants were observed only in WES+IMP. The coding variation was even more similar when limiting comparison to the target capture regions (Supplementary Table 4).

**Supplementary Figure 1. Flowchart of analytical UKB sample.** The analysis includes individuals from the UK Biobank with WES, imputed array, and WGS data with all analytical datasets in the dotted box. The primary analytical dataset includes 149,195 individuals who have all data sources available (bold); secondary datasets include (a) 468,169 individuals with WES and imputed array data and (b) a subset of 47,545 individuals with WGS.

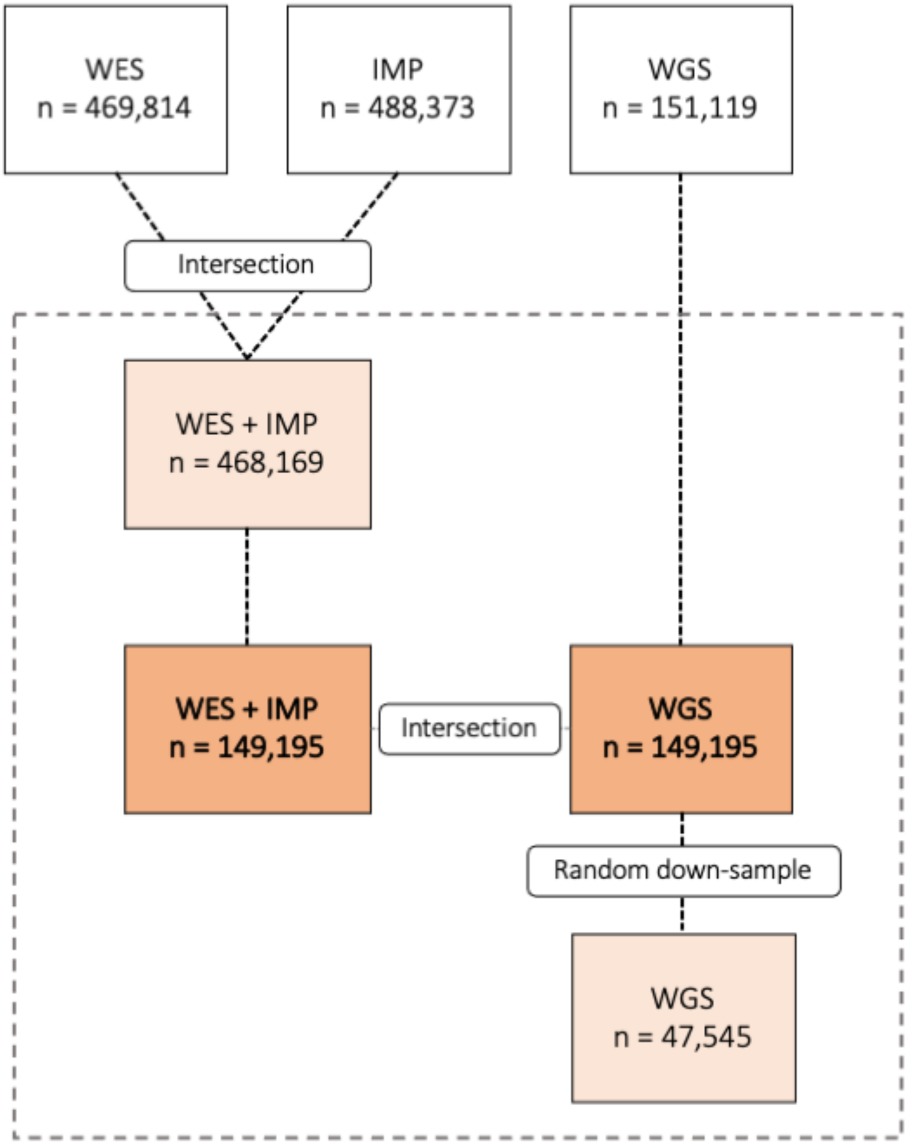

**Supplementary Figure 2. Flowchart of primary analyses.** Analyses of the primary datasets (n=149,195) included performing single variant and gene-based association testing for WES, imputed array, and WGS data. The same tests were performed on the secondary datasets (n=468,169 and n=47,545).

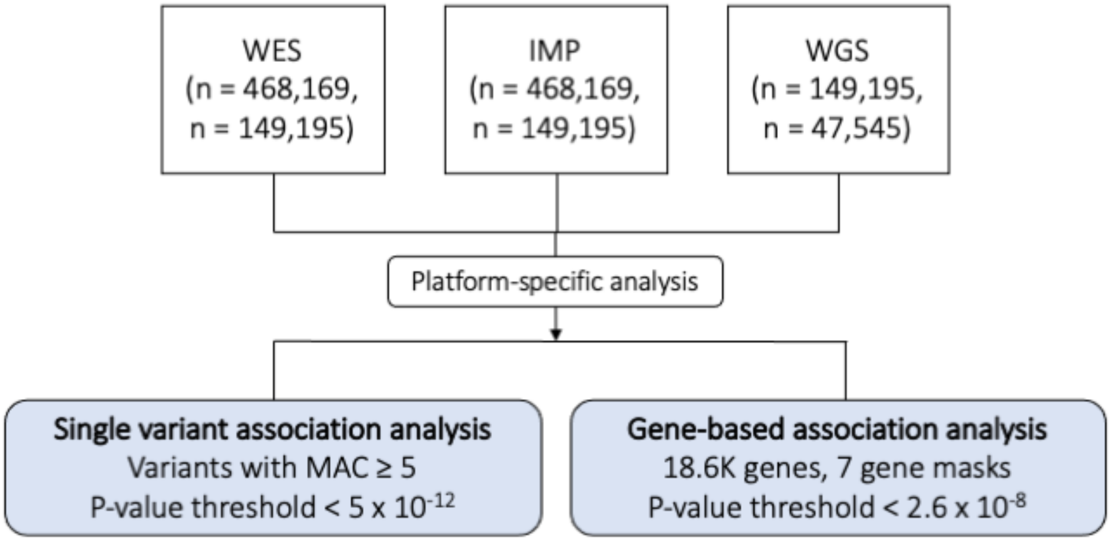

**Supplementary Figure 3. Survey of coding variation for WGS and WES+IMP.** A comparison of the coding variation observed by the WES+IMP and WGS datasets stratified by variant consequence. In Panel A, the count of variants observed in each approach, in both approaches, and in only one approach is given; the percentage gains in approach-specific variants is also given. In Panel B, the variant count per individual is given.

**A**

| Consequence | WGS (% Singleton) | WES+IMP (% Singleton) | Intersection | WGS only | WES+IMP only | % WGS only | % WES+IMP only |
|---|---|---|---|---|---|---|---|
| **Coding variants** | 6,732,108 (48%) | 6,761,880 (48%) | 6,544,263 | 187,845 | 217,617 | 2.7 | 3.1 |
| **Missense** | 4,225,468 (49%) | 4,263,339 (48%) | 4,137,020 | 88,448 | 126,319 | 2.0 | 2.9 |
| **Synonymous** | 1,994,972 (44%) | 2,012,866 (44%) | 1,959,714 | 35,258 | 53,152 | 1.7 | 2.6 |
| **In-frame indel** | 87,080 (51%) | 70,543 (45%) | 65,383 | 21,697 | 5,160 | 23.5 | 5.6 |
| **pLoF** | 424,588 (60%) | 415,132 (58%) | 382,146 | 42,442 | 32,986 | 9.3 | 7.2 |
| Frameshift | 190,713 (63%) | 182,447 (60%) | 162,815 | 27,898 | 19,632 | 13.3 | 9.3 |
| Stop gained | 130,968 (56%) | 132,452 (56%) | 126,845 | 4,123 | 5,607 | 3.0 | 4.1 |
| Start lost | 12,058 (52%) | 11,744 (51%) | 11,374 | 684 | 370 | 5.5 | 3.0 |
| Stop lost | 4,599 (58%) | 4,417 (57%) | 4,184 | 415 | 233 | 8.6 | 4.8 |
| Splice donor | 49,170 (60%) | 46,658 (59%) | 43,151 | 6,019 | 3,507 | 11.4 | 6.7 |
| Splice acceptor | 37,080 (62%) | 37,414 (61%) | 33,777 | 3,303 | 3,637 | 8.1 | 8.9 |

**B**

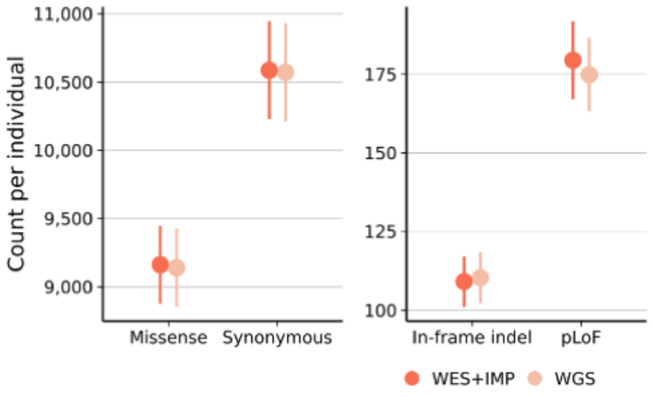

**Supplementary Figure 4. Flowchart of unified gene-p test.** Gene-based association analyses primarily focused on a single, unified p-value per gene. This gene-p p-value aggregates across multiple variant frequencies, masks (Supplementary Table 6), and set-based testing methods. The flowchart visualized how the single variants from a given gene are combined and tested to yield a single gene-level p-value, where gray arrows indicate aggregation by ACAT (Liu et al., 2019).

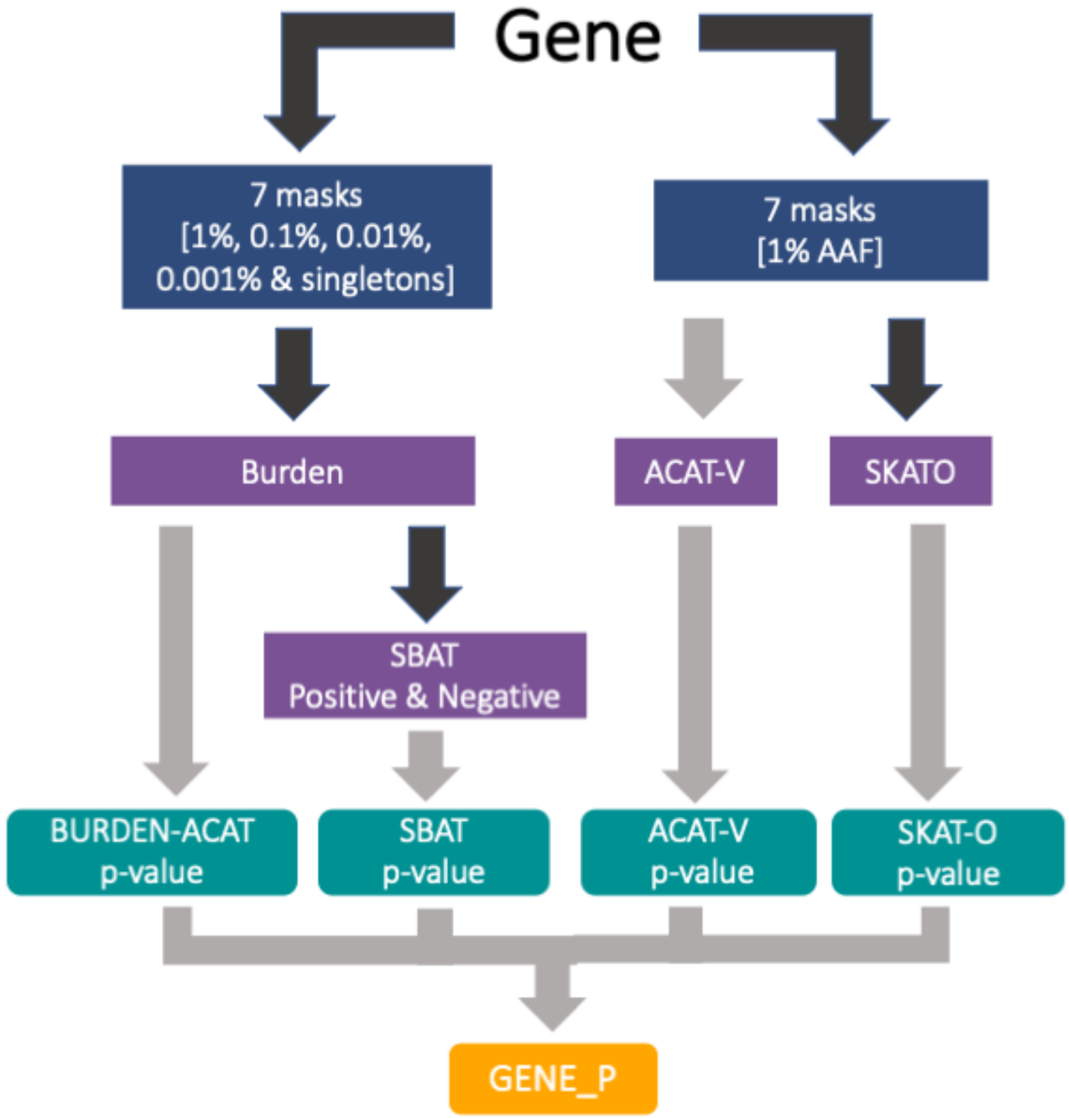

**Supplementary Figure 5. Summary of allele frequency, effect size, and p-value of all single variant associati on signals.** Single variant associations are grouped by the platform in which they were observed. Key features o f the signals, AAF, effect size, and -log10(p-value), are plotted for each signal in all groups.

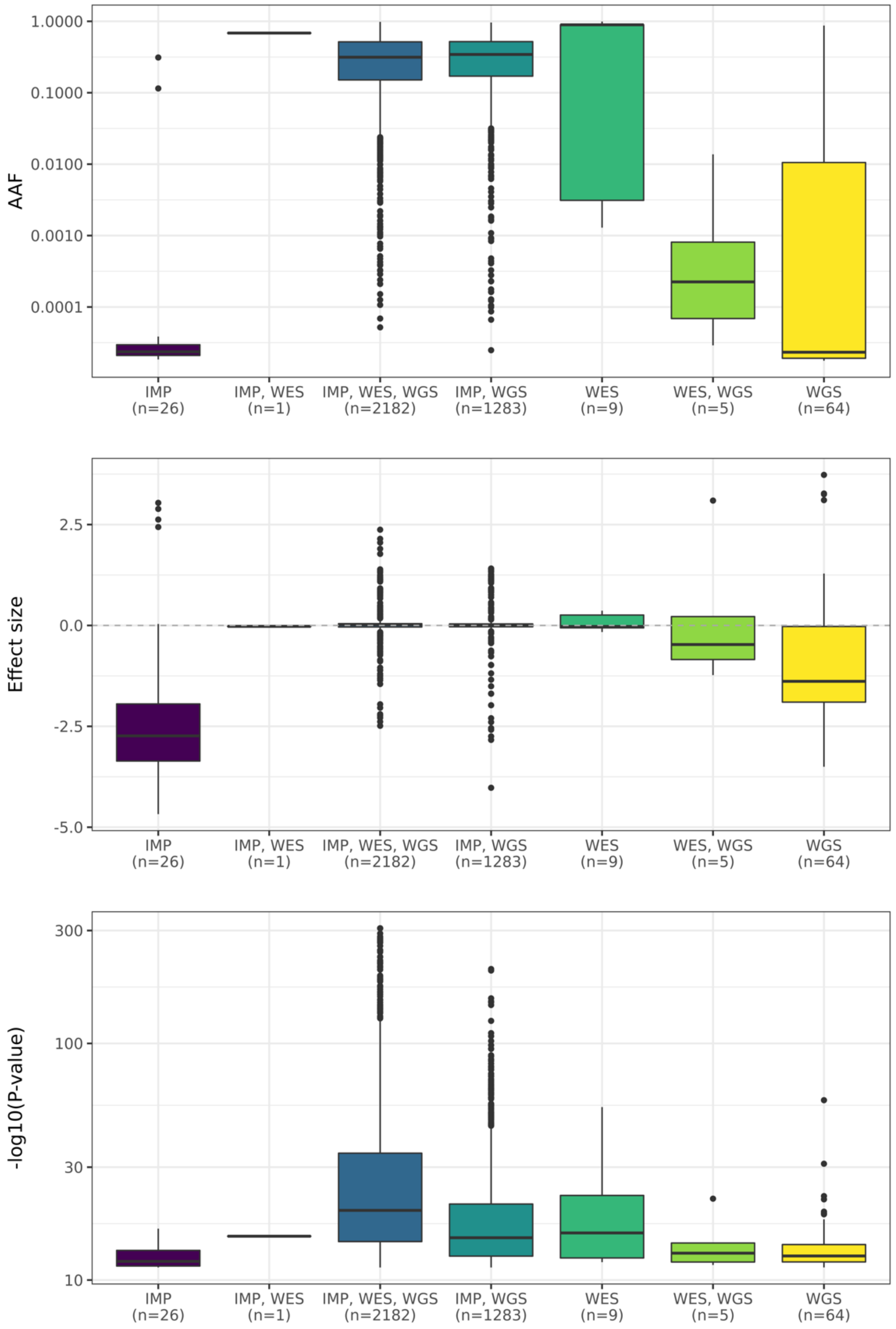

**Supplementary Figure 6. LocusZoom plot of lead single variant signals detected by all platforms.** A 1Mb region centered around most significant single variant association that was supported across all platforms (WGS, WES, IMP, WGS – SV). This was defined as observing an association with a p-value within an order of magnitude of the threshold of significance within a 1Mb region of the index association. The association is shown for 12:21178615:T:C (rs4149056), associated with Total bilirubin and identified first from the WGS sequencing data with p-value 2.23e-307. This missense variant lies in an exon of gene *SLCO1B1*, and is commonly observed with AAF=0.15.

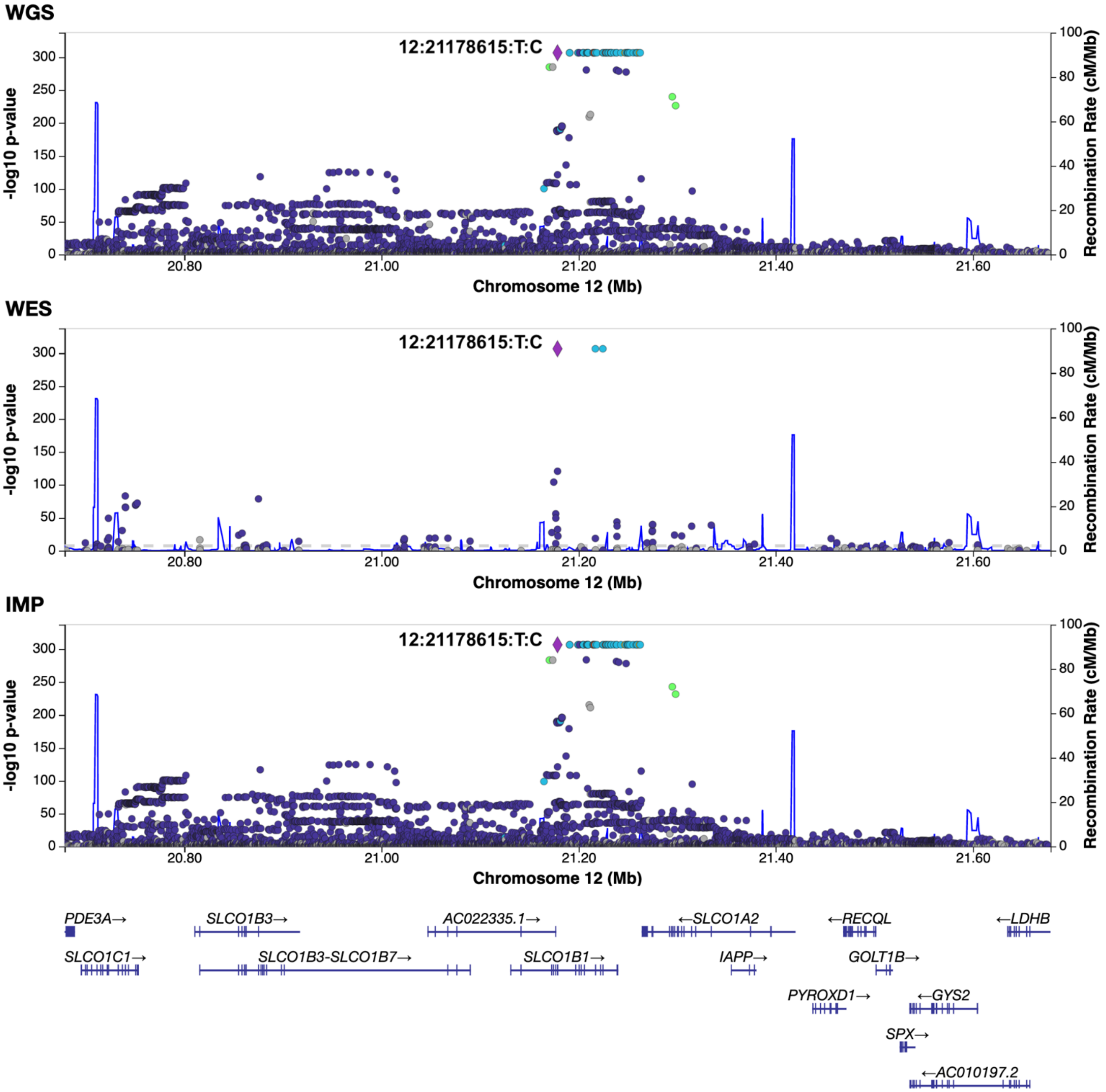

**Supplementary Figure 7. LocusZoom plots of single variant signals detected only by WGS.** A 1Mb region centered on a peak single variant association signal observed only in WGS. This variant, 7:73890306:C:T, is associated with standing height with p-value 4.67e-12 and AAF=0.13. It is supported by additional associated variants in WGS; the region also indicates support but below the 'suggestive' threshold for the same variant in the imputed data. The intergenic variant is not observed in the WES data.

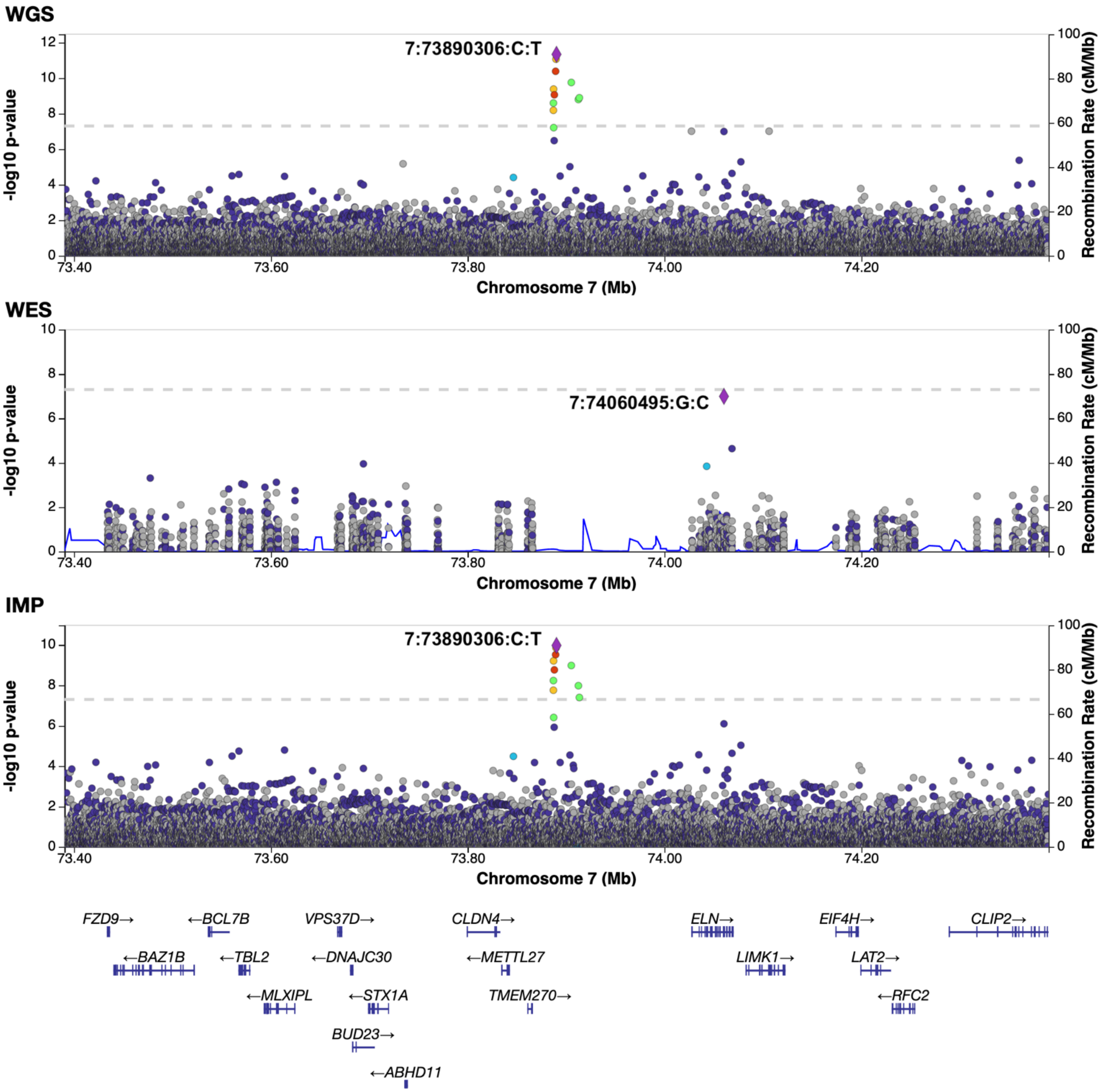

**Supplementary Figure 8. LocusZoom plots of single variant signals detected only by WES.** A 1Mb region cent ered on a peak single variant association signal observed only in WES. Variant 14:33367284:AAAG:A is an introni c variant n gene *NPAS3*. It is associated with mean reticulocyte volume with p-value 3.99e-13 and AAF 0.0020. T he most significant variant in the region in WES is neighboring with a p-value below the commonly recognized 5 e-8 GWAS threshold. Similar signal is not observed in WGS and IMP.

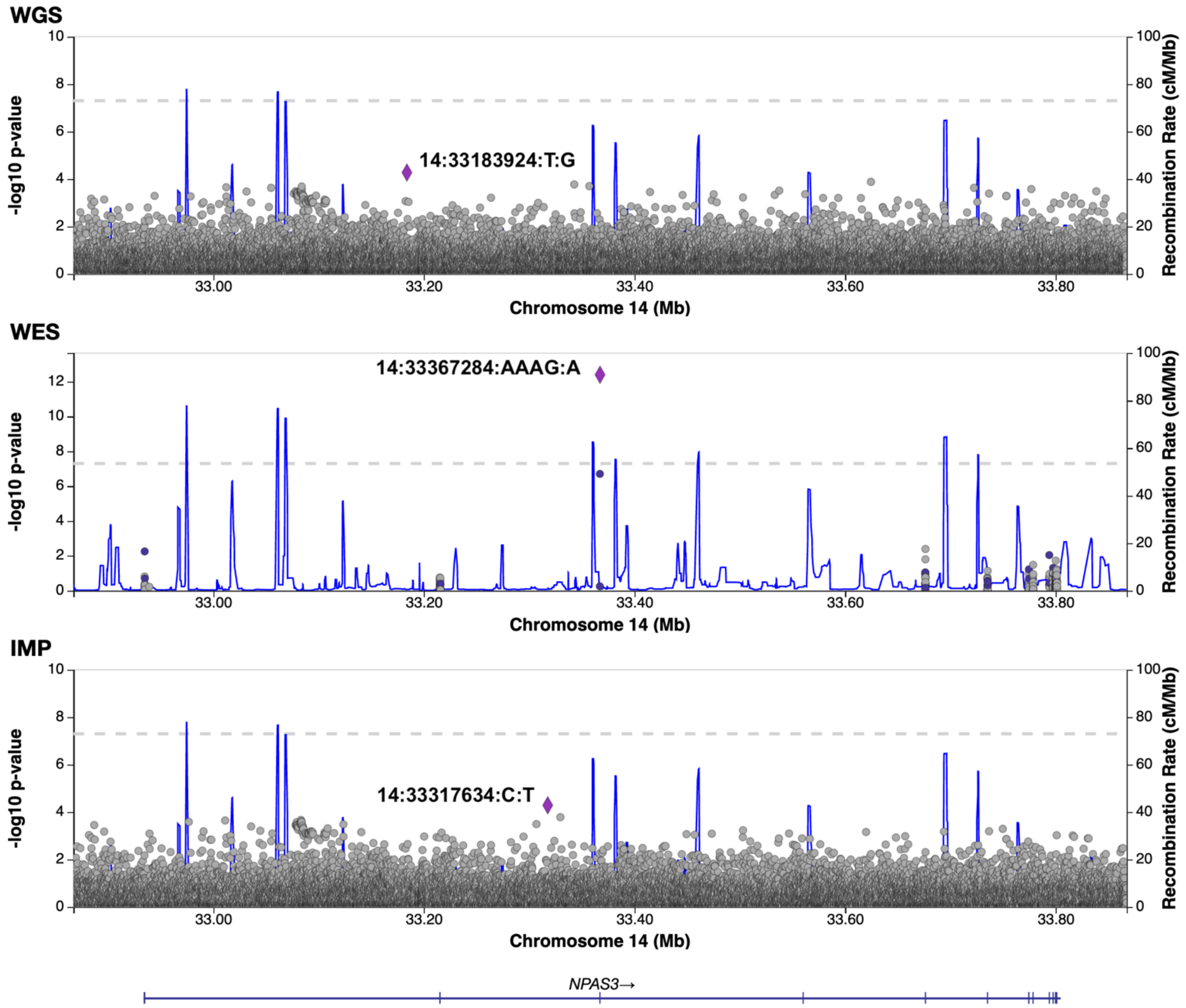

**Supplementary Figure 9. LocusZoom plots of single variant signals detected only by IMP.** A 1Mb region cent ered on a peak single variant association signal observed only in IMP. This intronic variant, 19:51628767:CGT:C, is associated with eosinophil count with p-value 4.37e-12 and AAF=0.31. It is supported by other variants in IMP , and the same peak variant and neighboring variants lie below the suggestive signal threshold in WGS.

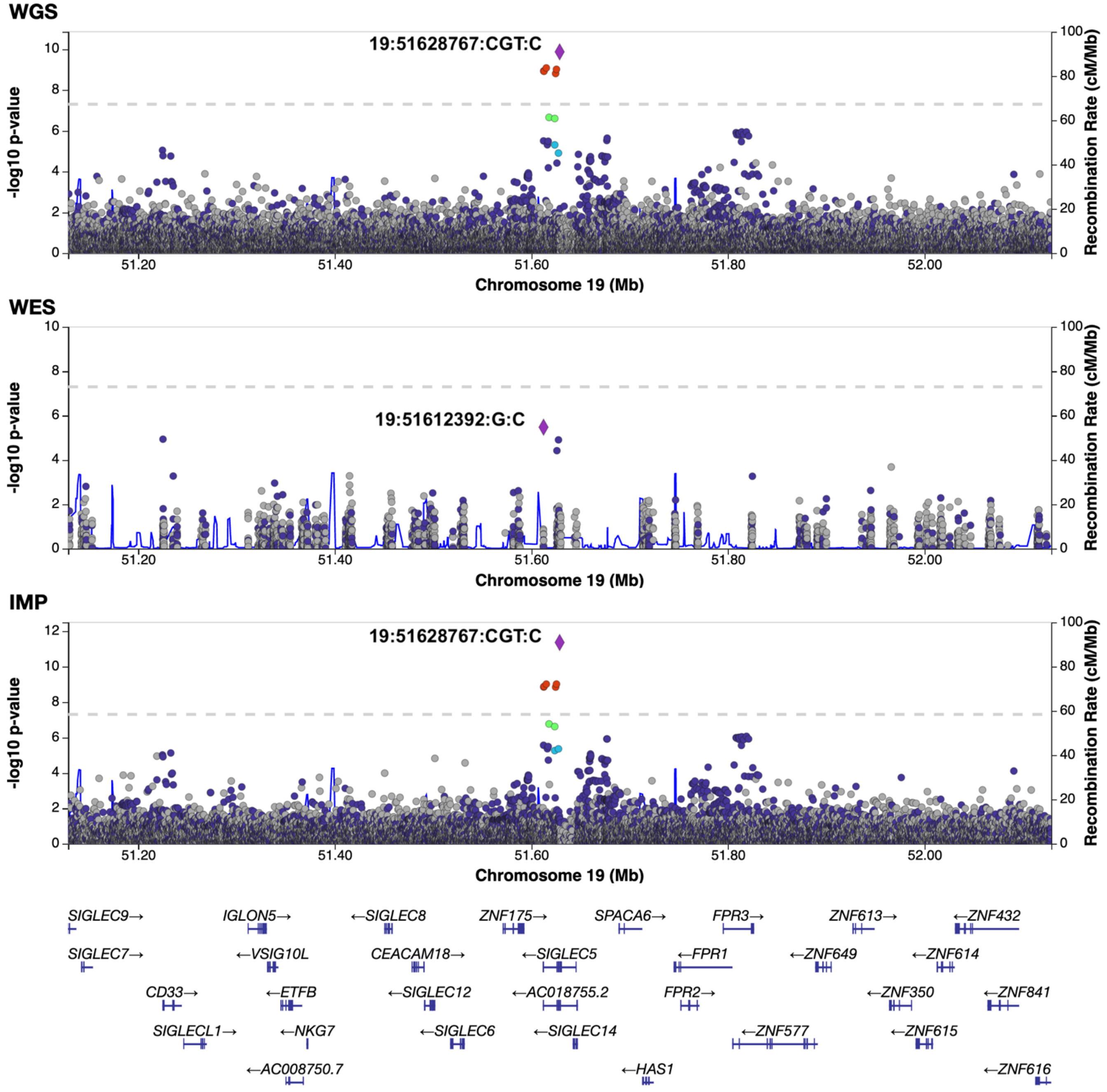

**Supplementary Figure 10. Summary of p-value of all gene-based association signals.** Gene-based associations are grouped by the platform in which they were observed. The unified gene P p-value, incorporating multiple statistical tests and masks, are plotted for each signal in all groups.

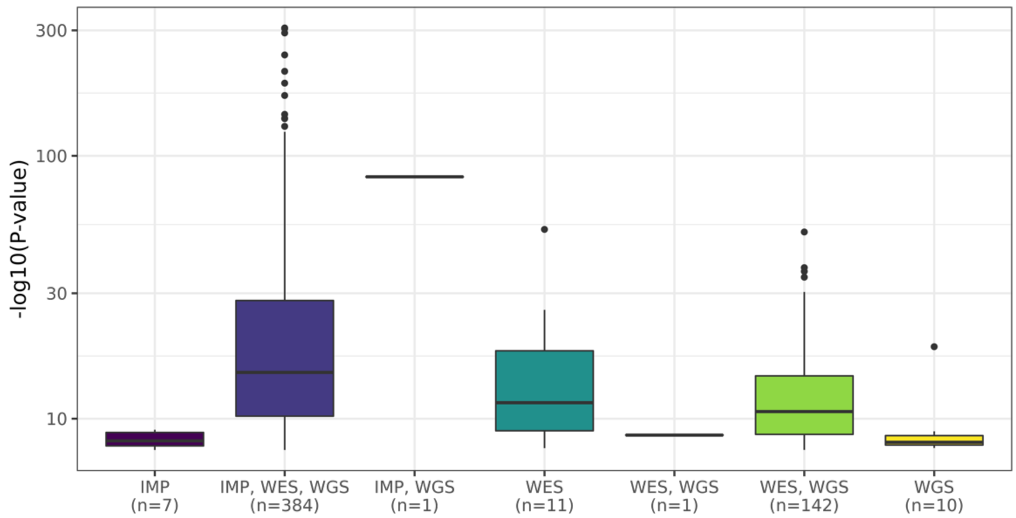

**Supplementary Figure 11. Comparison of gene-p p-values for gene-based analyses between platforms.** For each gene tested, the p-value between each pair of platforms is given for all tests and for those with $-\log_{10}(p - value)$.

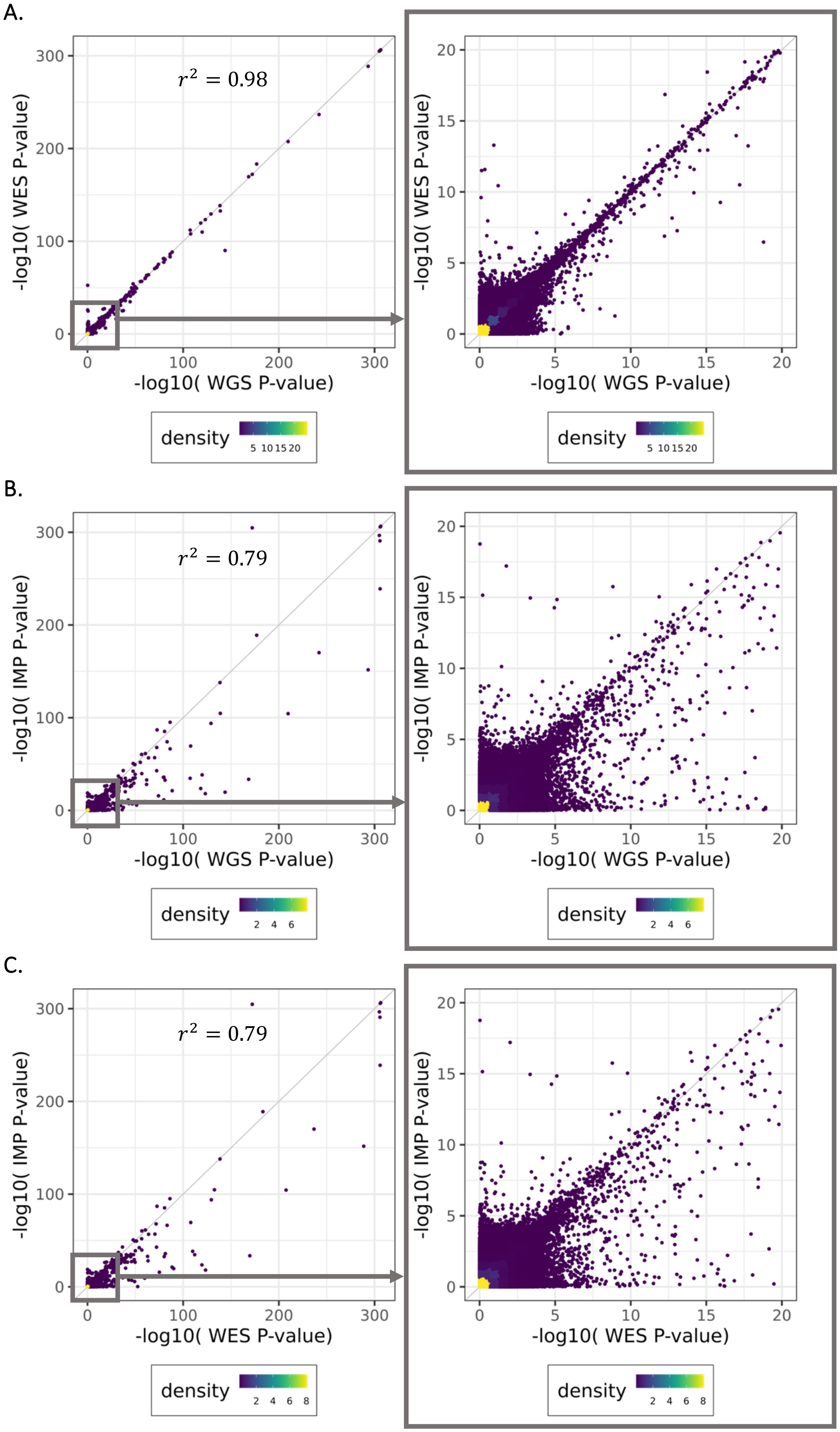

**Supplementary Table 1. Characteristics of the UKB data.** Analyses included UKB data with three sample sizes, comprised of individuals with an assigned ancestry. Demographic features of these individuals are provided.

| Sample size | Platforms analyzed | Sex - Female | Mean age (SD) | Ancestry | |
|---|---|---|---|---|---|
| 47,545 | WGS | 26,384 (55.5%) | 56.5 (8.1) | 982 | AFR |
| | | | | 97 | AMR |
| | | | | 227 | EAS |
| | | | | 45,093 | EUR |
| | | | | 1,146 | SAS |
| 149,195 | IMP, WES, WGS | 82,210 (55.1%) | 56.5 (8.1) | 3,008 | AFR |
| | | | | 277 | AMR |
| | | | | 736 | EAS |
| | | | | 141,695 | EUR |
| | | | | 3,479 | SAS |
| 468,169 | IMP, WES | 253,697 (54.2%) | 56.5 (8.1) | 9,277 | AFR |
| | | | | 856 | AMR |
| | | | | 2,303 | EAS |
| | | | | 445,544 | EUR |
| | | | | 10,189 | SAS |

**Supplementary Table 2. Genotype concordance between the different approaches.** For all autosomal variants that passed QC in each platform, after enforcing hard calls, we assessed the number of mean discordant calls per variant and the concordance across all variant calls.

| Datasets | Number of variants compared | Mean number of discordant calls per variant | Concordance |
|---|---|---|---|
| WGS & WES | 15,840,174 | 9.6 | 99.99% |
| WES & IMP | 2,371,567 | 75.3 | 99.95% |
| WGS & IMP | 89,809,894 | 72.2 | 99.95% |

**Supplementary Table 3. Number of canonical coding variants in WGS and WES+IMP datasets.** Count of variants for each coding consequence, stratified by frequency. Variants were annotated with VEP and genes were defined by Ensembl v100.

| Consequence | Frequency | WGS | WES+IMP | Union | Intersection | WGS only | WES+IMP only |
|---|---|---|---|---|---|---|---|
| Missense | Singleton (AAC=1) | 2,055,463 | 2,056,701 | 2,113,288 | 1,998,876 | 56,587 | 57,825 |
| | AAC>1 & AAF<=0.0001 | 1,935,023 | 1,968,371 | 1,999,707 | 1,903,687 | 31,336 | 64,684 |
| | AAF>0.001 & AAF<=0.01 | 208,238 | 211,273 | 211,740 | 207,771 | 467 | 3,502 |
| | AAF>0.01 | 26,744 | 26,994 | 27,052 | 26,686 | 58 | 308 |
| Stop gained | Singleton (AAC=1) | 73,515 | 73,664 | 76,502 | 70,677 | 2,838 | 2,987 |
| | AAC>1 & AAF<=0.0001 | 53,823 | 55,045 | 56,312 | 52,556 | 1,267 | 2,489 |
| | AAF>0.001 & AAF<=0.01 | 3,429 | 3,537 | 3,554 | 3,412 | 17 | 125 |
| | AAC>1 & AAF<=0.0001 | 53,823 | 55,045 | 56,312 | 52,556 | 1,267 | 2,489 |
| | AAF>0.01 | 201 | 206 | 207 | 200 | 1 | 6 |
| Synonymous | Singleton (AAC=1) | 884,307 | 886,016 | 907,833 | 862,490 | 21,817 | 23,526 |
| | AAC>1 & AAF<=0.0001 | 943,930 | 958,636 | 971,785 | 930,781 | 13,149 | 27,855 |
| | AAF>0.001 & AAF<=0.01 | 138,548 | 139,882 | 140,132 | 138,298 | 250 | 1,584 |
| | AAF>0.01 | 28,187 | 28,332 | 28,374 | 28,145 | 42 | 187 |
| In-frame indel | Singleton (AAC=1) | 44,268 | 31,537 | 46,642 | 29,163 | 15,105 | 2,374 |
| | AAC>1 & AAF<=0.0001 | 38,109 | 34,143 | 40,573 | 31,679 | 6,430 | 2,464 |

| | | | | | | | |
|---|---|---|---|---|---|---|---|
| | AAF>0.001 & AAF<=0.01 | 4,311 | 4,450 | 4,599 | 4,162 | 149 | 288 |
| | AAF>0.01 | 392 | 413 | 426 | 379 | 13 | 34 |
| Frameshift | Singleton (AAC=1) | 120,430 | 108,817 | 130,794 | 98,453 | 21,977 | 10,364 |
| | AAC>1 & AAF<=0.0001 | 65,982 | 68,518 | 74,362 | 60,138 | 5,844 | 8,380 |
| | AAF>0.001 & AAF<=0.01 | 4,065 | 4,813 | 4,878 | 4,000 | 65 | 813 |
| | AAF>0.01 | 236 | 299 | 311 | 224 | 12 | 75 |
| Splice donor | Singleton (AAC=1) | 27,177 | 27,000 | 29,192 | 24,985 | 2,192 | 2,015 |
| | AAC>1 & AAF<=0.0001 | 16,549 | 17,209 | 17,875 | 15,883 | 666 | 1,326 |
| | AAF>0.001 & AAF<=0.01 | 1,097 | 1,143 | 1,155 | 1,085 | 12 | 58 |
| | AAF>0.01 | 63 | 67 | 70 | 60 | 3 | 7 |
| Splice acceptor | Singleton (AAC=1) | 21,344 | 21,693 | 23,321 | 19,716 | 1,628 | 1,977 |
| | AAC>1 & AAF<=0.0001 | 11,993 | 12,814 | 13,320 | 11,487 | 506 | 1,327 |
| | AAF>0.001 & AAF<=0.01 | 721 | 749 | 752 | 718 | 3 | 31 |
| | AAF>0.01 | 39 | 41 | 41 | 39 | 0 | 2 |
| Start lost | Singleton (AAC=1) | 6,250 | 5,968 | 6,418 | 5,800 | 450 | 168 |
| | AAC>1 & AAF<=0.0001 | 5,351 | 5,307 | 5,537 | 5,121 | 230 | 186 |
| | AAF>0.001 & AAF<=0.01 | 421 | 432 | 436 | 417 | 4 | 15 |
| | AAF>0.01 | 36 | 37 | 37 | 36 | 0 | 1 |
| Stop lost | Singleton (AAC=1) | 2,678 | 2,509 | 2,810 | 2,377 | 301 | 132 |
| | AAC>1 & AAF<=0.0001 | 1,751 | 1,736 | 1,847 | 1,640 | 111 | 96 |
| | AAF>0.001 & AAF<=0.01 | 153 | 155 | 158 | 150 | 3 | 5 |
| | AAF>0.01 | 17 | 17 | 17 | 17 | 0 | 0 |
| 5’ UTR splice acceptor | Singleton (AAC=1) | 1,559 | 1,113 | 1,741 | 931 | 628 | 182 |
| | AAC>1 & AAF<=0.0001 | 1,066 | 811 | 1,171 | 706 | 360 | 105 |
| | AAF>0.001 & AAF<=0.01 | 94 | 98 | 99 | 93 | 1 | 5 |
| | AAF>0.01 | 10 | 11 | 11 | 10 | 0 | 1 |
| 3’ UTR splice acceptor | Singleton (AAC=1) | 134 | 23 | 136 | 21 | 113 | 2 |
| | AAC>1 & AAF<=0.0001 | 103 | 42 | 106 | 39 | 64 | 3 |
| | AAF>0.001 & AAF<=0.01 | 16 | 18 | 18 | 16 | 0 | 2 |
| | AAF>0.01 | 1 | 1 | 1 | 1 | 0 | 0 |
| 5’ UTR splice donor | Singleton (AAC=1) | 2,119 | 284 | 2,139 | 264 | 1,855 | 20 |
| | AAC>1 & AAF<=0.0001 | 1,655 | 476 | 1,699 | 432 | 1,223 | 44 |
| | AAF>0.001 & AAF<=0.01 | 155 | 152 | 158 | 149 | 6 | 3 |
| | AAF>0.01 | 25 | 25 | 25 | 25 | 0 | 0 |
| 3’ UTR splice donor | Singleton (AAC=1) | 178 | 153 | 199 | 132 | 46 | 21 |
| | AAC>1 & AAF<=0.0001 | 124 | 121 | 136 | 109 | 15 | 12 |

| | | | | | | | |
|---|---|---|---|---|---|---|---|
| | AAF>0.001 & AAF<=0.01 | 20 | 21 | 21 | 20 | 0 | 1 |
| | AAF>0.01 | 8 | 7 | 8 | 7 | 1 | 0 |

**Supplementary Table 4. Number of canonical coding variants in target capture regions for WGS and WES+IMP datasets.** Count of variants for each coding consequence when limiting to variants within the WES targeted capture regions. Variants were annotated with VEP and genes were defined by Ensembl v100. pLoFs included frameshift, splice donor, splice acceptor, stop gained, stop lost, and start lost variants.

| Consequence | WGS | WES+IMP | Union | Intersection | WGS only | WES+IMP only |
|---|---|---|---|---|---|---|
| Missense | 4155578 | 4244803 | 4279583 | 4120798 | 34780 | 124005 |
| Synonymous | 1964215 | 2003872 | 2016349 | 1951738 | 12477 | 52134 |
| In-frame indel | 84619 | 69754 | 89655 | 64718 | 19901 | 5036 |
| pLoF | 358211 | 356825 | 385531 | 329505 | 28706 | 27320 |

**Supplementary Table 5. Single variant signal consequences by platforms with association observed.** Count of variants for each consequence with an observed trait association, given by the platforms in which the signal is observed. Variants were annotated with VEP and genes were defined by Ensembl v100.

| Consequence | IMP | IMP, WES | IMP, WES, WGS | IMP, WGS | WES | WES, WGS | WGS |
|---|---|---|---|---|---|---|---|
| 3' UTR | 0 | 0 | 75 | 27 | 0 | 0 | 1 |
| 5' UTR | 0 | 0 | 27 | 9 | 0 | 0 | 0 |
| Downstream | 1 | 0 | 108 | 23 | 0 | 0 | 2 |
| Frameshift | 0 | 0 | 5 | 0 | 0 | 1 | 0 |
| In-frame indel | 0 | 0 | 6 | 0 | 0 | 0 | 0 |
| Intergenic | 17 | 0 | 422 | 691 | 0 | 0 | 38 |
| Intronic | 8 | 1 | 1,014 | 467 | 7 | 2 | 22 |
| Missense | 0 | 0 | 281 | 0 | 1 | 1 | 0 |
| Splice donor | 0 | 0 | 2 | 0 | 0 | 1 | 0 |
| Splice region | 0 | 0 | 6 | 0 | 0 | 0 | 0 |
| Stop gained | 0 | 0 | 10 | 0 | 0 | 0 | 0 |
| Synonymous | 0 | 0 | 28 | 0 | 0 | 0 | 0 |
| Upstream | 0 | 0 | 198 | 66 | 1 | 0 | 1 |

**Supplementary Table 6. Gene burden mask definitions.** For gene-based testing, variants were grouped into seven different masks by variant consequence. The variants were annotated with VEP and Ensembl 100, and aggregated into masks for tests in Regenie.

| Mask | Variant consequences included |
|---|---|
| pLoF | stop_gained, stop_gain, frameshift, splice_donor, splice_acceptor |
| pLoF_missense_5 | stop_gained, stop_gain, frameshift, splice_donor, splice_acceptor, start_lost, stop_lost, missense(5/5) |
| pLoF_missense_1 | stop_gained, stop_gain, frameshift, splice_donor, splice_acceptor, start_lost, stop_lost, missense(5/5), missense(>=1/5), UTR_splice_donor, 5_prime_UTR_splice_donor, 3_prime_UTR_splice_donor, UTR_splice_acceptor, 5_prime_UTR_splice_acceptor, 3_prime_UTR_splice_acceptor |
| pLoF_missense_0 | stop_gained, stop_gain, frameshift, splice_donor, splice_acceptor, start_lost, stop_lost, missense(5/5), missense(>=1/5), missense(0/5), UTR_splice_donor, |

| | |
|---|---|
| | 5_prime_UTR_splice_donor, 3_prime_UTR_splice_donor, UTR_splice_acceptor, 5_prime_UTR_splice_acceptor, 3_prime_UTR_splice_acceptor |
| missense_5 | start_lost, stop_lost, missense(5/5) |
| missense_1 | start_lost, stop_lost, missense(5/5), missense(>=1/5) |
| missense_0 | start_lost, stop_lost, missense(5/5), missense(>=1/5), missense(0/5) |

### Supplementary Table 7. Significant single variant tests.
*Attached*.

### Supplementary Table 8. Significant gene-based tests.
*Attached*.
